# Saliva is a promising alternative specimen for the detection of SARS-CoV-2 in children and adults

**DOI:** 10.1101/2020.10.25.20219055

**Authors:** Rebecca Yee, Thao Truong, Pia S. Pannaraj, Natalie Eubanks, Emily Gai, Jaycee Jumarang, Lauren Turner, Ariana Peralta, Yesun Lee, Jennifer Dien Bard

## Abstract

Testing efforts for SARS-CoV-2 have been burdened by the scarcity of testing materials and personal protective equipment for healthcare workers. The simple and painless process of saliva collection allows for widespread testing, but enthusiasm is hampered by variable performance compared to nasopharyngeal swab (NPS) samples. We prospectively collected paired NPS and saliva samples from a total of 300 unique adult and pediatric patients. SARS-CoV-2 RNA was detected in 32.2% (97/300) of the individuals using the TaqPath COVID-19 Combo Kit (Thermo Fisher). Performance of saliva and NPS were compared against the total number of positives regardless of specimen type. The overall concordance for saliva and NPS was 91.0% (273/300) and 94.7% (284/300), respectively. The positive percent agreement (PPA) for saliva and NPS was 81.4% (79/97) and 89.7% (87/97), respectively. Saliva detected 10 positive cases that were negative by NPS. In symptomatic and asymptomatic pediatric patients not previously diagnosed with COVID-19, the performances of saliva and NPS were comparable (PPA: 82.4% vs 85.3%). The overall PPA for adults were 83.3% and 90.7% for saliva and NPS, respectively, with saliva detecting 4 cases less than NPS. However, saliva performance in symptomatic adults was identical to NPS (PPA of 93.8%). With lower cost and self-collection capabilities, saliva can be an appropriate alternative sample choice to NPS for detection of SARS-CoV-2 in children and adults.

**Summary:** Saliva is an acceptable alternative specimen compared to nasopharyngeal swabs for detection of SARS-CoV-2. Specifically, saliva demonstrated comparable performance to nasopharyngeal swabs in symptomatic and asymptomatic pediatric patients and in symptomatic adults.

## Introduction

Accurate and timely molecular testing for SARS-CoV-2, the causative agent of the ongoing coronavirus disease 2019 (COVID-19) pandemic, has been crucial for informing patient management, public health decision making, contact tracing, and infection control. The Infectious Diseases Society of America (IDSA) guidelines recommend testing for SARS-CoV-2 by reverse-transcriptase polymerase chain reaction (RT-PCR) on specimen samples which includes nasopharyngeal swabs (NPS), mid-turbinate swabs, or nasal swabs rather than oropharyngeal swabs (OPS) or saliva alone (1). However, testing efforts have been hampered by supply chain shortages due to an unprecedented demand for testing materials such as swabs, universal transport media, and personal protective equipment for healthcare workers (2). The simplicity of saliva collection has certainly increased its interest as an alternative specimen for detection of SARS-CoV-2.

Compared to NP specimen collection, saliva is less invasive, circumvents the need for swabs, and requires minimal supervision with the option for self-collection. Previous studies have indicated that saliva is a promising specimen for detection of other respiratory viruses by RT-PCR, including influenza and common non-SARS human coronaviruses (3-5). To-date, the U.S. Food and Drug Administration has issued several emergency use authorizations for laboratory-developed diagnostic tests using saliva. More recent studies have shown use of saliva has moderate-to-high sensitivity and specificity when compared to NP swabs for detection of SARS-CoV-2 (6-12). These studies vary widely in sample collection method and testing platforms, and more data is needed to standardize best collection and processing practices.

There is tremendous motivation to pursue saliva collection in children, not only because of the simplicity in specimen collection but to also avoid the unnecessary discomfort during NPS collection. There is also huge interest in saliva as a primary specimen type to detected SARS-CoV-2 during the school year. Hence, it is important to understand the dynamics of viral detection in children, which has implications for their contribution to transmission of SARS-CoV-2. Unfortunately, data on the use of saliva to detect SARS-CoV-2 in pediatric patients is sparse. The few reports available on the performance of saliva specimens in children showed poor detection of SARS-CoV-2 with sensitivities of 53-73% albeit such studies suffer from small sample sizes (13-15). In this study, we evaluated and compared prospectively collected paired saliva and NP swabs from both pediatric and adult patients for detection of SARS-CoV-2. We also compare the differences in viral load in asymptomatic and symptomatic COVID-19 patients.

## Methods

### Study Design

A total of 300 unique patients (inpatients, outpatients and household members of diagnosed COVID-19 patients) were enrolled in this study between June 8 to August 28, 2020. Demographic data including age, gender, and symptoms were collected. Participants were asked if they had previously tested positive for COVID-19. Paired samples were collected from individuals with unknown COVID-19 status as well as from patients previously positive for SARS-CoV-2. Both symptomatic and asymptomatic patients were enrolled in the study. Study design conducted at Children’s Hospital Los Angeles was approved by the Institutional Review Board under IRB #CHLA-20-00124 and CHLA-18-00098.

### Sample collection

At least 3 mL of saliva was self-collected under the observation of a healthcare worker who subsequently collected a NP swab sample for parallel testing. Saliva was collected in a sterile cup and NP swabs were immediately placed in viral transport medium (Becton Dickenson, Franklin Lakes, NJ, USA). Samples were either sent to the clinical laboratory within 1 hour from collection or stored at 4°C and sent to the clinical laboratory within 4 hours from collection. Samples were stored at 4°C and tested within 48 hours from collection or stored at -80°C prior to testing.

### qRT–PCR assay for SARS-CoV-2 RNA

Paired nasopharyngeal swabs and saliva were sent to the Clinical Virology Laboratory at Children’s Hospital Los Angeles. Total nucleic acid was extracted from 250 µL samples using the Thermo Fisher KingFisher Flex specimen processing system with the Applied Biosystems MagMAX Viral/Pathogen Nucleic Acid Isolation Kit (Thermo Fisher, Waltham, MA) and eluted to 50 µL of total nucleic acid. Real-time reverse transcription polymerase chain reaction (RT-PCR) was performed using the TaqPath COVID-19 Combo Kit (Thermo Fisher). A positive result for SARS-CoV-2 detection was determined by amplification of at least one of the three genes targeted (N gene, S gene or ORF1ab gene) using a cut-off of Ct value <40. A valid negative result for SARS-CoV-2 detection was determined by amplification of MS2 internal control using a cut-off of Ct value <32.

### Data and Statistical analysis

A composite gold standard approach was used to determine a true positive case. Any positive detected from either NPS or saliva was considered a true positive and positive percent agreement (PPA) and negative percent agreement (NPA) was calculated based on this. Statistical analyses comparing different Ct values and days between onset of symptoms and test date were performed using a Mann-Whitney test.

## Results

During a 11-week period (June 8 to August 28, 2020), SARS-CoV-2 RNA was detected in a total of 97 out of 300 individuals, of which 43 (44.3%) were < 19 years of age. The median age was 37.5 years old (range 19-58) and 12 years old (range 4-18) in our adult and pediatric COVID-19 positive cohorts, respectively. A female predominance was noted (61/97, 62.9%). Of the 97 COVID-19 positive patients, 55 (56.7 %) were symptomatic at the time of collection with a median of 10 days between symptom onset and time of collection. Twenty-seven (27.8%) patients were known to be positive for SARS-CoV-2 prior to enrollment. Since individuals in entire households were enrolled, it was not surprising that an overwhelming proportion of our cohort (73/97, 75.3%) reported exposure to a COVID-19 positive individual.

The overall concordance of saliva and NPS was 91.0% (273/300) and 94.7% (284/300), respectively. When analyzing all 97 positive patients, SARS-CoV-2 RNA were detected from both NPS and saliva in 69 patients, from saliva only in 10 patients and NPS only in 18 patients. The overall PPA for saliva and NPS was 81.4% (79/97) and 89.7% (87/97), respectively, when compared to a total number of positive cases identified by RT-PCR (Table 1). The NPA was 100% for both specimen types.

**Table 1.**
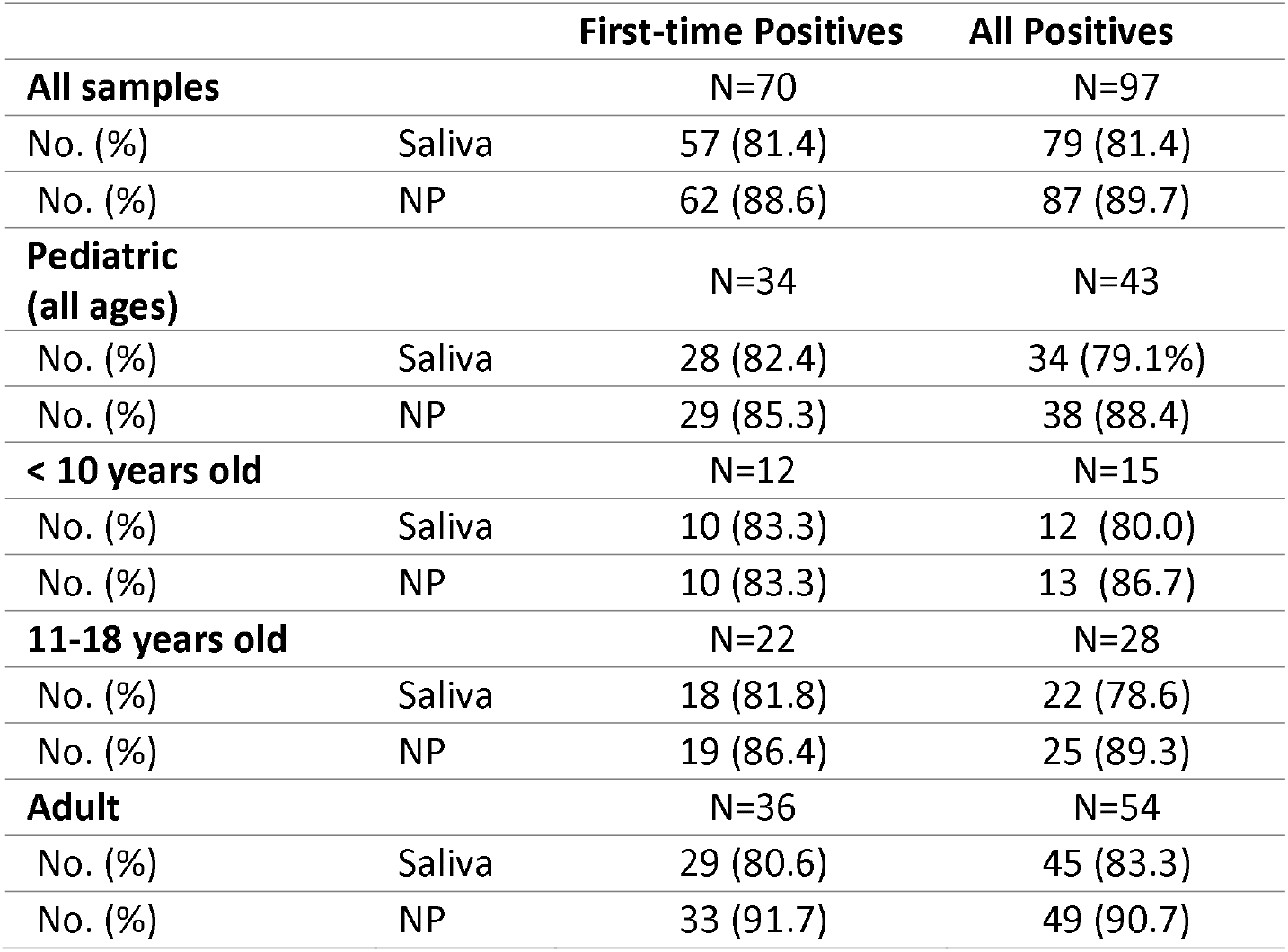
Performance of Saliva and NP specimens.

|  |  | First-time Positives | All Positives |
| --- | --- | --- | --- |
| **All samples** |  | N=70 | N=97 |
| No. (%) | Saliva | 57 (81.4) | 79 (81.4) |
| No. (%) | NP | 62 (88.6) | 87 (89.7) |
| **Pediatric (all ages)** |  | N=34 | N=43 |
| No. (%) | Saliva | 28 (82.4) | 34 (79.1%) |
| No. (%) | NP | 29 (85.3) | 38 (88.4) |
| **< 10 years old** |  | N=12 | N=15 |
| No. (%) | Saliva | 10 (83.3) | 12 (80.0) |
| No. (%) | NP | 10 (83.3) | 13 (86.7) |
| **11-18 years old** |  | N=22 | N=28 |
| No. (%) | Saliva | 18 (81.8) | 22 (78.6) |
| No. (%) | NP | 19 (86.4) | 25 (89.3) |
| **Adult** |  | N=36 | N=54 |
| No. (%) | Saliva | 29 (80.6) | 45 (83.3) |
| No. (%) | NP | 33 (91.7) | 49 (90.7) |

Focusing on pediatric patients only, the overall PPA were 79.1% for saliva and 88.4% for NPS collected. Performance of saliva (PPA: 82.4%) and NPS (PPA: 85.3%) were comparable when only first time positive pediatric patients were analyzed for both symptomatic and asymptomatic patients. Specifically, testing using saliva detected the same number of COVID-19 cases as NPS (both at 78.6%) in the asymptomatic pediatric cohort and only missed one positive case (85% vs 90%) in the symptomatic cohort (Table 2). The performance of saliva remained high in both young and older children. In children ages 4-10 years, saliva and NPS achieved PPA of 83.3%. Additionally, saliva was able to capture all 6/6 (100%) symptomatic patients in this age group as opposed to the 5/6 (83.3%) for NPS. In older patients between 11-18 years old, one positive case was missed by saliva (PPA: 81.8% vs 86.4%) but the performance was superior when testing only asymptomatic patients (PPA: 87.5% vs 75.0%) with detection of an additional case (Table 2).

**Table 2.** Performance of Saliva and NP specimens in Symptomatic Patients.

|  |  | Symptomatic (%) |  | Asymptomatic (%) |  |
| --- | --- | --- | --- | --- | --- |
|  |  | First-time Positives | All Positives | First-time Positives | All Positives |
| **All samples** |  | N=38 | N=55 | N=32 | N=42 |
| No. (%) | Saliva | 34 (89.5) | 49 (89.1) | 23 (71.9) | 30 (71.4) |
| No. (%) | NP | 36 (94.7) | 51 (92.7) | 26 (81.3) | 36 (85.7) |
| **All Pediatric (0-18 y)** |  | N=20 | N=23 | N=14 | N=20 |
| No. (%) | Saliva | 17 (85.0) | 19 (82.6) | 11 (78.6) | 15 (75.0) |
| No. (%) | NP | 18 (90.0) | 21 (91.3) | 11 (78.6) | 17 (85.0) |
| **< 10 y** |  | N=6 | N=8 | N=6 | N=7 |
| No. (%) | Saliva | 6 (100) | 7 (87.5) | 4 (66.7) | 5 (71.4) |
| No. (%) | NP | 5 (83.3) | 7 (87.5) | 5 (83.3) | 6 (85.7) |
| **11-18 y** |  | N=14 | N=15 | N=8 | N=13 |
| No. (%) | Saliva | 11 (78.6) | 12 (80.0) | 7 (87.5) | 10 (76.9) |
| No. (%) | NP | 13 (92.9) | 14 (93.3) | 6 (75.0) | 11 (84.6) |
| **Adult (>18 y)** |  | N=18 | N=32 | N=18 | N=22 |
| No. (%) | Saliva | 17 (94.4) | 30 (93.8) | 12 (66.7) | 15 (68.2) |
| No. (%) | NP | 18 (100) | 30 (93.8) | 15 (83.3) | 19 (86.4) |

In adult patients, the overall PPA were 83.3% and 90.7% for saliva and NPS, respectively. In contrast to the pediatric data, saliva performed better in symptomatic patients with identical PPA to NPS at 93.8% but poorly in asymptomatic adults (PPA: 68.2% vs 86.4). Findings were comparable even when only first time positive patients were analyzed. (Table 1-2).

The average differences in Ct values between saliva and NPS samples were not statistically different (Ct: 28.7 versus 29.1) (Figure 1A-B). Based on linear regression analysis where Ct values of saliva (Y-axis) are plotted against the Ct values of NPS (X-axis) from the paired sample, the equation of y=0.9994x suggests that Ct values from both sample types are approximately equivalent to one another (Figure 1C). In addition, the Ct values of both saliva and NPS samples remain comparable regardless of age and disease status (symptomatic vs asymptomatic) (Figure 2).

**Figure 1.**
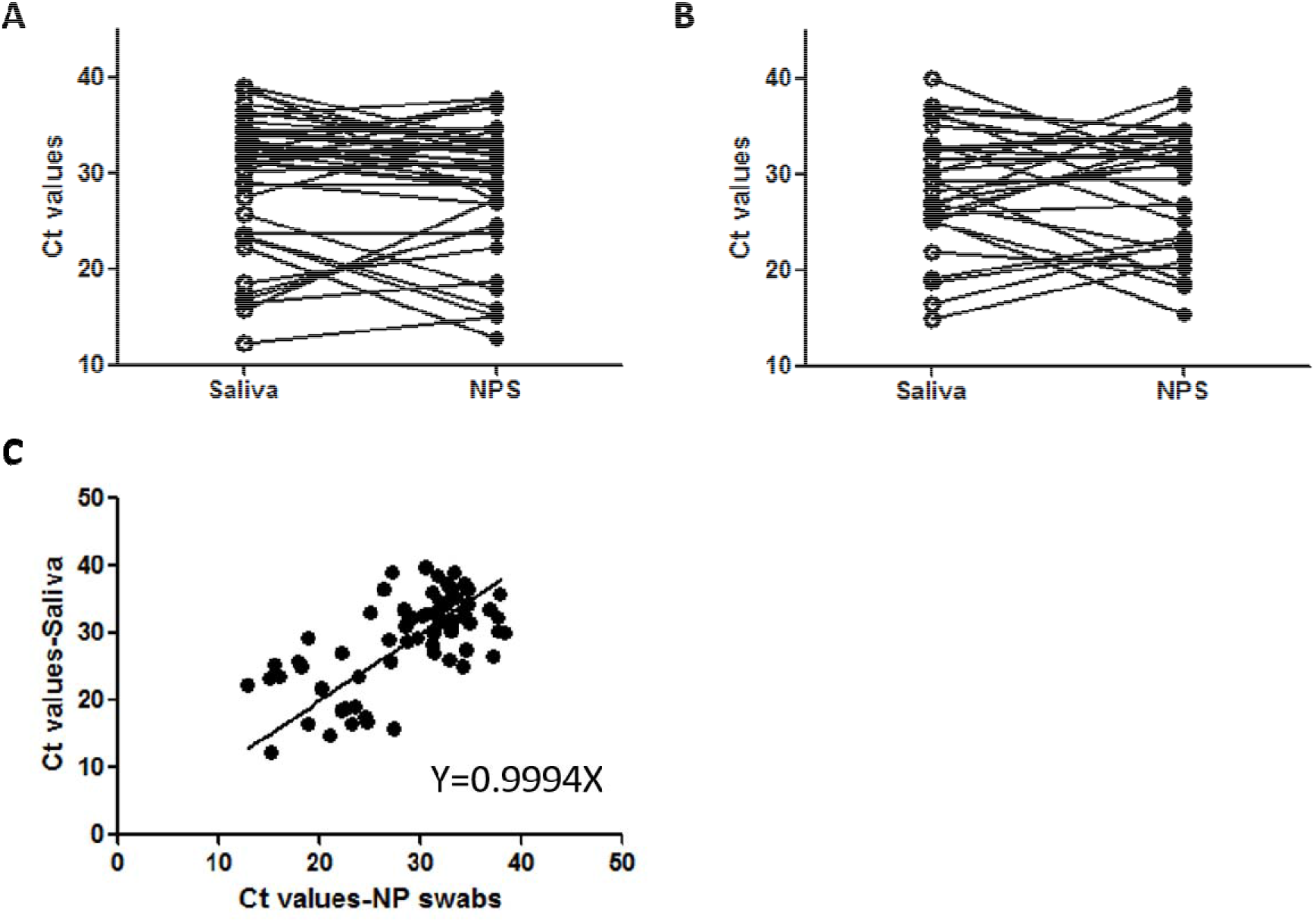
Concordance of Ct values from Saliva and NP swabs. Comparison of Ct values from paired saliva and nasopharyngeal swab specimens in (A) adult and (B) pediatric patients that were positive for SARS-CoV-2. Each line represents the corresponding paired specimen. (C) Regression curve plotting Ct values from paired saliva and nasopharyngeal swab specimens that were positive for SARS-CoV-2 reveal a linear association between the Ct values obtained from the two specimen types.

**Figure 2.**
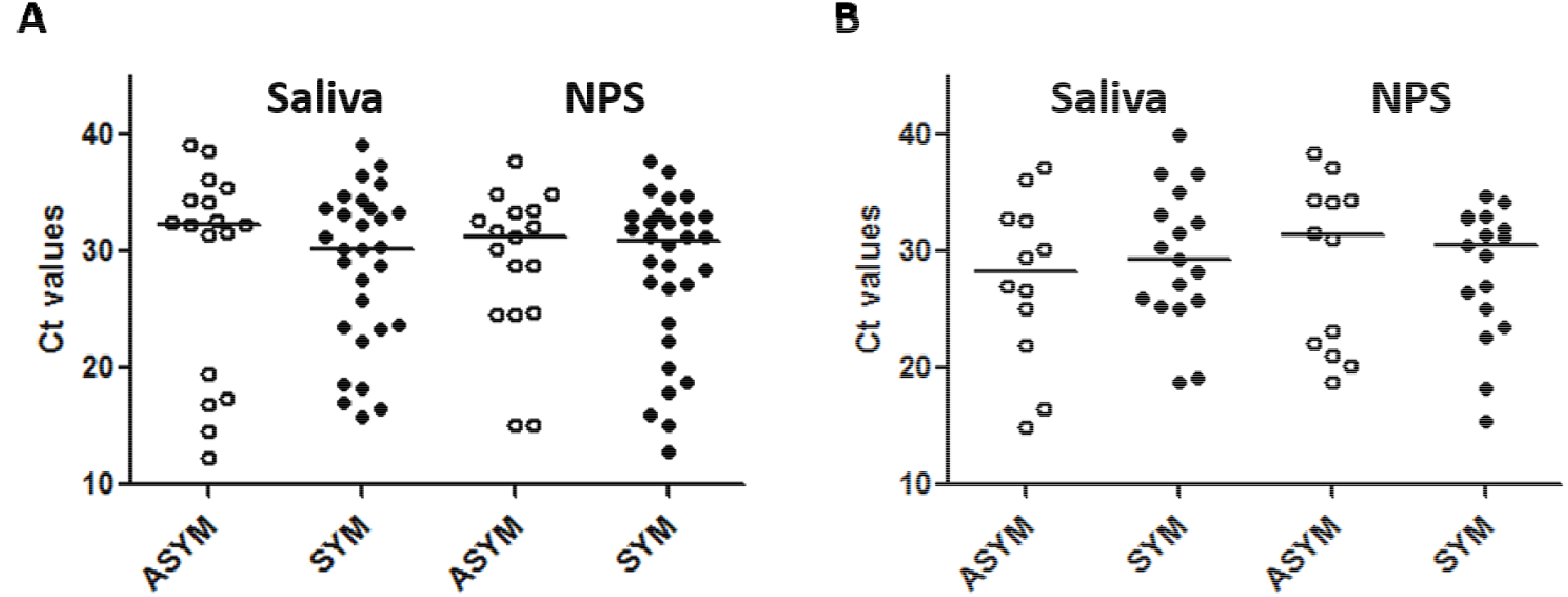
Comparison of Ct values from asymptomatic and symptomatic populations. The Ct values from saliva and nasopharyngeal swab specimens collected from our SARS-CoV-2 positive asymptomatic (open circle) and symptomatic (filled circle) patients in our (A) adult populations and (B) pediatric cohort.

Importantly, SARS-CoV-2 RNA were detected in 28 (28.9%) patients in only one sample type (10 saliva; 18 NPS). Most of these patients were older than 10 years (25/28, 89.3%) (Supplementary Table 1). Saliva-only positive patients were tested ranging from 3 to 43 days post-symptom onset compared to the 7 to 31 day post-symptom onset in NPS-only positive patients. The overall Ct values between saliva-only and NPS-only positives were comparable (Ct of 32.4 vs 32.5) with 88.8% (NPS-positive only) and 80% (saliva-positive only) of the samples having a Ct of over 30 (Figure 3).

**Figure 3.**
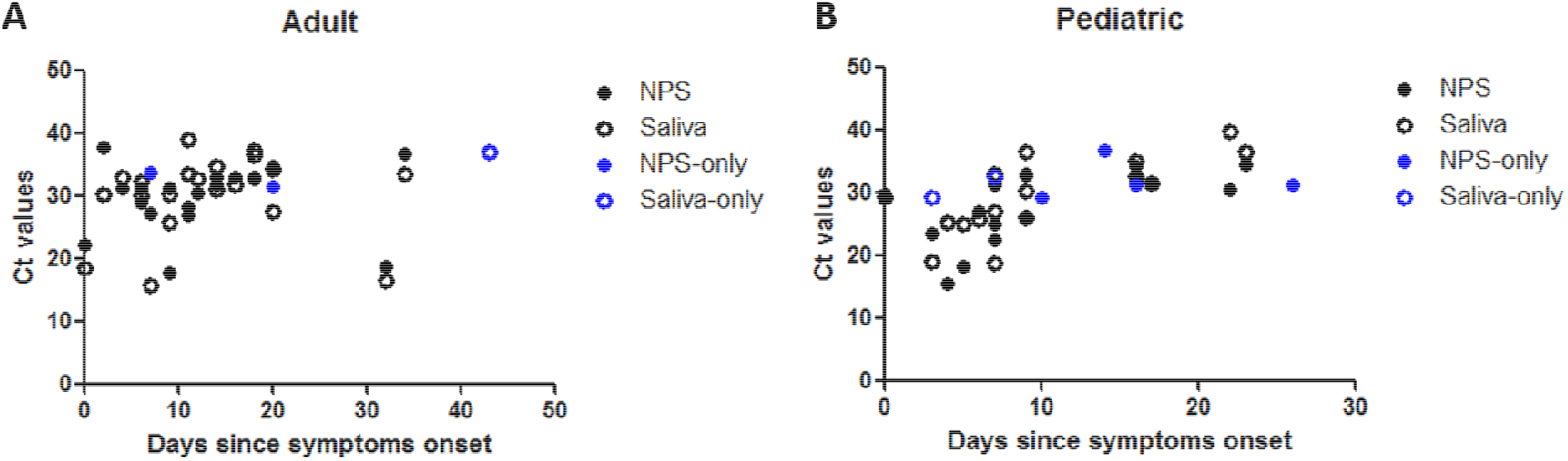
Ct values of saliva and NP swab samples in relation to days between time of symptom onset to time of collection for testing. The Ct values of (A) adult and (B) pediatric patients tested positive by both nasopharyngeal swab (black solid circle) and saliva (black open circle), nasopharyngeal swab only (blue filled circle), and saliva only (blue open circle) are depicted in reference to when they were tested since symptom onset (days).

The average Ct values derived from cases detected by both saliva and NPS was lower than when only one sample type was positive (Ct 28.9 vs Ct 32.4, p<0.001). Symptomatic patients were more likely to have SARS-CoV-2 RNA detected from both sample types (OR=3.37, p=0.01).

## Discussion

Testing saliva specimens can circumvent the shortage of collection supplies and may be a sufficient noninvasive and more cost-effective alternative for SARS-CoV-2 testing (4). The sensitivity of saliva for detection of SARS-CoV-2 has been shown to be less than NPS in other studies, ranging from 72% to 86% (16, 17). We demonstrated an overall PPA of 81.4% in saliva versus 89.7% in NPS in our entire cohort. Comparable performance of saliva to NPS was shown in children who were previously unknown positive patients (both symptomatic and asymptomatic patients) and also in symptomatic adults only. To our knowledge, this is the first and largest study demonstrating support for utilization of saliva in the pediatric age group and comparison of performance of saliva between pediatric and adult cohorts.

It is important to note that testing of saliva caught 10 additional COVID-19 cases that were negative by NPS. Our findings are consistent with results from other studies demonstrating how saliva specimens can identify otherwise missed cases of not only COVID-19, but also influenza and RSV (4, 6, 17). In this study, of the 18 samples that were detected by NPS only, 7 (38.9%) were from asymptomatic adults, a subpopulation that performed poorly with detection of SARS-CoV-2 in saliva. Additionally, over 80% of NPS-positive only patients exhibited Ct values past 30.0, suggesting that false negatives are attributed to lower viral loads. Additionally, our study showed that the performance of saliva is not dependent on age which is corroborated by recent studies which also reported that age had no impact on viral load and detection of SARS-CoV-2 (15, 18), including in pediatric populations.

While some studies argue that viral load is highest in saliva within the first week of symptom onset, others have shown that saliva can be more sensitive than NPS throughout the course of infection or sometimes produce intermittent positive results over the course of a few weeks (19). A small, longitudinal pediatric study from South Korea found SARS-CoV-2 RNA was more readily detected from saliva within the first few days of symptom onset followed by a drastic decline in viral load compared to NPS (14). In contrast, we report the detection of SARS-CoV-2 in saliva for up to 43 days compared to 32 days for NP swabs.

While several studies have shown that NPS yield lower Ct values than saliva in symptomatic adult patients (8, 10, 11), we report no significant difference in Ct values between saliva and NPS in either our adult or pediatric patients. Our findings corroborates with a recent study of 19 adults that reported no significant differences (7). Interestingly, a recent study demonstrated that in adult populations, performance of saliva was better than NPS in detecting SARS-CoV-2 in asymptomatic individuals, but our results suggest that saliva was a poor alternative to NPS in asymptomatic adults, missing 4 cases that were NPS positive (20). However, it must be noted that in our older children cohort (11-18 years old), saliva’s performance was superior than NPS for detection in first-time positive asymptomatic individuals. The conflicting findings between studies may be due to differences in saliva collection protocol, collection device, age of patient, and also the inherent difficulties in working with a more viscous sample that may be more prone to more sampling variabilities (9, 10). Such differences in methodology may account for the variability in the performance of saliva reported in other studies. A more thorough comparison and standardization of saliva collection and processing needs to be evaluated.

Limitations of this study include the small sample size of both children, particularly younger children, and adults from a single medical institution. Second, this study consisted of only outpatients, patients admitted to the emergency department, and family members who volunteered to enroll in the study which can bias our findings regarding the role of COVID-19 exposure to specimen performance. Since viral load may or may not be correlated with clinical manifestations, further studies should be conducted in inpatient or ICU settings as the spectrum of disease ranges from asymptomatic to severely ill patients (21-23). Finally, despite a standardized protocol utilized during the collection of the saliva samples, it can be challenging for children to properly salivate into a collection device. The volume of saliva obtained may also vary among patients due to excessive bubbles and other factors despite the same amount of saliva being processed for testing.

## Conclusions

Our study reveals that saliva is a reliable diagnostic specimen for the detection of SARS-CoV-2 RNA by RT-PCR, particularly in both symptomatic and asymptomatic children and symptomatic adults. Moreover, saliva was able to identify additional COVID-19 cases that were otherwise missed by NPS. With saliva collection being a more cost-effective and non-invasive approach, it offers a feasible approach for widespread testing of SARS-CoV-2 in the inpatient settings and in the community.

## Data Availability

The manuscript contains all relevant data.

## Acknowledgments

We would like to acknowledge the staff members of the Clinical Virology laboratory at Children’s Hospital Los Angeles for dedication towards SARS-CoV-2 RT-PCR testing. We would like to acknowledge Dr. Javier Mestas and Irvin Ibarra Flores for assisting with the saliva testing.

