## Supplemental Table 1 for "Saliva is a promising alternative specimen for the detection of SARS-CoV-2 in children and adults"

### Supplementary Table

#### Supplementary Table 1. Characteristics of patients with only one specimen type positive for SARS-CoV-2

| | | Saliva (N=10) | NP swab (N=18) |
|---|---|---|---|
| **Sex** | | | |
| | Female | 8 (80.0) | 13 (72.2) |
| | Male | 2 (20.0) | 5 (27.8) |
| **Age** | | | |
| | Median, range (years) | | |
| | 1-5 | 0 (0) | 0 (0) |
| | 5-10 | 1 (10.0) | 2 (11.1) |
| | 10-18 | 4 (40.0) | 7 (38.9) |
| | 18-30 | 1 (10.0) | 2 (11.1) |
| | 30-40 | 0 (0) | 3 (16.7) |
| | 40-50 | 4 (40.0) | 3 (16.7) |
| | 50-60 | 0 (0) | 1 (5.6) |
| **COVID-19 exposure** | | | |
| | No | 5 (50.0) | 2 (11.1) |
| | Yes | 5 (50.0) | 16 (88.9) |
| **Symptomatic** | | | |
| | No | 5 (50.0) | 12 (66.7) |
| | Yes | 5 (50.0) | 6 (33.3) |
| **Time of symptom onset from date tested** | | | |
| | Day (range) | 13.5 (2-43) | 15 (7-26) |
